# Basic Reproduction Rate and Case Fatality Rate of COVID-19: Application of Meta-analysis

**DOI:** 10.1101/2020.05.13.20100750

**Authors:** Suryakant Yadav, Pawan Kumar Yadav

**Affiliations:** Assistant Professor, Department of Development Studies, International Institute for Population Sciences, Mumbai. (;); Research Scholar, International Institute for Population Sciences, Mumbai.

**Keywords:** Covid-19, SARS-CoV-2, Reproduction Rate, Case Fatality Rate, Systematic Review, Meta-analysis

## Abstract

**Background:** The outbreak of novel coronavirus disease of 2019 (COVID-19) has a wider geographical spread than other previous viruses such as Ebola and H1N1. The onset of disease and its transmission and severity has become a global concern. The policymakers have a serious concern for containing the spread and minimising the risk of death.

**Aim:** This study aims to provide the estimates of basic reproduction rate (R_0_) and case fatality rate (CFR) which applies to a generalised population.

**Methods:** A systematic review was carried out to retrieve the published estimates of reproduction rate and case fatality rate in peer-reviewed articles from PubMed MEDLINE database with defined inclusion and exclusion criteria in the period 15 December 2019 to 3 May 2020. The systematic review led to the selection of 24 articles for R_0_ and 17 articles for CFR. These studies used data from China and its provinces, other Asian countries such as Japan, Korea, the Philippines, and countries from other parts of the world such as Nigeria, Iran, Italy, Europe as a whole, France, Latin America, Turkey, the United Kingdom (UK), and the United States of America (USA). These selected articles gave an output of 30 counts of R_0_ and 29 counts of CFR which were used in a meta-analysis. A meta-analysis, with the inverse variance method, fixed- and random-effects model and the Forest plot, was performed to estimate the mean effect size or mean value of basic reproduction rate and case fatality rate. The Funnel plot is used to comprehend the publication bias.

**Results:** We estimated the robust estimate of R_0_ at 3.11 (2.49–3.71) persons and the robust estimate of CFR at 2.56 (2.06–3.05) per cent after accounting for heterogeneity among studies, using the random-effects model. The regional subgroup analysis in a meta-analysis was significant for R_0_ but was not significant for CFR. The R_0_ values varied from 1.90 (1.06–2.74) persons to 3.83 (2.44–5.22) persons across the regions. The Funnel plot confirms that the selected studies are significant at one per cent level of significance.

**Conclusion:** We found that one person is likely to infect two to three persons in the absence of any control measures, and around three per cent of the population are at the risk of death within one-and-a-half months from the onset of disease COVID-19 in a generalised population. The emergence of SARS-CoV-2 varies across regions, but the risk of death remains the same.

**Contribution:** The estimates of R_0_ and CFR are independent of data from a particular region or time or a homogeneous population. These estimates are applicable to a generalised population. Therefore, the estimates of R_0_ and CFR are unequivocally applicable to developing country like India and its states or districts, in ambivalence. The assessments of R_0_ and CFR values across the developed nations make all of us aware of consequences of COVID-19, and hence these estimates are of crucial importance for government authorities for the practical implementation of strategies and control measures to contain the disease.

Research Highlights

1. The robust estimate of Basic Reproduction Rate (R_0_) of COVID-19 based on a meta-analysis performed on the pieces of evidence available across countries is 3.11 (2.49–3.71) persons for a generalised population in the absence of any control measures
2. The robust estimate of Case Fatality Rate (CFR) based on a meta-analysis performed on the pieces of evidence available across countries equals to 2.56 (2.06–3.05) per cent for a generalised population in approximately one-and-a-half months from the onset of the disease COVID-19.
3. A significant regional variation is evident for the Basic Reproduction Rate (R_0_) but not for the Case Fatality Rate (CFR)
4. The peer-reviewed articles with a small sample size do not suffer from publication bias in a meta-analysis of COVID-19.

Added Value of this Study
Out study combine available evidence of the parameter values, such as reproduction rate and case fatality rate, of the generalised epidemiological models for coronavirus disease of 2019 (COVID-19). In this way, we have reduced the dependency on data from a particular region or time or a homogeneous population. By applying meta-analysis, we estimated the robust estimate of reproduction rate and case fatality rate, which is applicable across heterogeneous populations. We proclaim that the reproduction rate of COVID-19 varies across subgroups of populations and regions and periods, but the case fatality rate remained the same. These estimates of reproduction rate and case fatality rate are worthwhile for developing countries like India and at a lower level of geography, in ambivalence.

## Introduction

The outbreak and spread of novel coronavirus-2019, particularly identified as Severe Acute Respiratory Syndrome Coronavirus 2 (SARS-CoV-2), in Wuhan City, Hubei Province, China, was first time reported to World Health Organisation (WHO) on 31 December 2019. On 7 January 2020, it was confirmed by Chinese authorities, and on 12 January 2020, WHO confirmed it. The epidemics of Severe Acute Respiratory Syndrome (SARS) and Middle East Acute Respiratory Syndrome (MARS) in the past and the recent outbreaks of infectious diseases like Ebola and H1N1 influenza make us conscientious about comprehending the spread of such viruses and consequences regarding life losses, morbidity, economic burden, and political instability. It is utmost crucial to control the spread and outbreak at the very first case of the disease. It is highly transmittable zoonotic coronavirus disease. The first step for academicians and any government agencies, beforehand, is to study the basic reproduction rate (R_0_) which is the average number of secondary infectious cases produced by an infectious case, that provides the plausibility of the spread, outbreak, and severity of an epidemic in a short time available to them. The value of R_0_ greater than one indicates the number of infectious persons is likely to increase and the value of R_0_ less than one indicates the transmission is likely to die out. R_0_ determines the potential for an epidemic spread in a susceptible population in the absence of specific control measures (Koff, 1992; World Health Organization, 2003). For understanding the transmissibility of SARS-CoV-2 in population, every country had made efforts for estimating the R_0_. Numerous studies have shown that recovery rate has remained more or less the same, but reproduction rate and case fatality rate (CFR) varied across the regions (Fanelli and Piazza 2020, p. 4). The case fatality rate is defined as the percentage of individuals with symptomatic or confirmed diseases who die from the disease. The case fatality rate is currently in the range of 2–8 per cent but has varied across the age groups – higher at old ages and lower at child and adult ages. The estimates of CFR based on hospital records lies in the range of 8–28 per cent (Verity et al. 2020, p. 2). These estimates are evidence to the severity of coronavirus in a short time of its outbreak.

The incubation period of R_0_ is in the range of 5 to 14 days which may be followed by death in without treatments or in no control measures. The severity of this disease remained suspicious because the onset of the disease is often unknown, and crude CFR is heavily underreported. Therefore, in the absence of control measures and no vaccination, this disease for an infected person is just fatal–the high immunity level to protect a person is an assumption in this case. The effect of MARS and SARS are known in the past; however, the variants of SARS or MARS are acute killer diseases.

The effective reproduction rate (R_t_), which is the potential for epidemic spread at a time ‘t’ under the control measures, is a function of R_0_ and proportion of the susceptible population (Cao et al., 2020). Since the incidence of new cases of SARS-CoV-2, the R_0_ is crucial to understand the mechanism as well as the implementation strategies to reach an effective value of R_t_ which should be less than one for containing the outbreak. The epidemic is considered to be under control when R_t_ is less than one. The CFR is quite easily measurable and less sensitive to censoring and bias compared to R_0_. The moments of the distribution of death and mortality patterns are most useful for calculating adjusted CFR; however, R_0_ is quite sensitive to the onset of diseases and the number of days to get a prediction. In other cases, it is R_t_ which is time-variant and mathematically, a limiting case of R_0_. Nevertheless, R_0_ is the most warranted statistics of the epidemiological model for COVID-19.

This study aims to provide, from a systematic review and a meta-analysis, a summary statistic of R_0_ and CFR which has the best applicability for other regions or countries as well as the lower level of geography districts and towns/village at the point of the onset of disease in a susceptible population. This summary statistics of R_0_ and CFR would be of immense utility to government authorities for the practical implementation of strategies or control measures at the very initial phase of the COVID-19 disease transmission.

## Objectives

1. To summarise the characteristics of studies specific to the basic reproduction rate (R_0_) and case fatality rate (CFR) of COVID-19
2. To access and compute the basic reproduction rate and case fatality rate among the confirmed cases of COVID-19

## Methods

We accessed the works of literature from the PubMed MEDLINE database. Based on the terms “coronavirus” in All Fields in PubMed MEDLINE search, the search in the database showed 3536 number of articles. With “COVID-19” in the field Title/Abstract, it showed 2248 number of articles. The preliminary basic search showed significant works of literature that are available to allow work on the systematic review of overall COVID-19.

### A systematic review of R_0_

From this broad set of works of literature, we selected research articles with description on reproduction number/rates, based on the terms ‘coronavirus reproduction’ in All Fields AND “COVID-19” in the field Title/Abstract. It gave an output of 173 number of articles. After applying filters for the English language, Free full text, and between the dates 15/12/2019 and 03/05/2020, it gave an output of 90 number of research articles which are peer-reviewed only for screening, so that we consider the complete works of literature. After reviewing the keywords and abstracts of these articles, we come to know that many of these works of literature have not worked on estimation of the parameters of epidemiological models such as reproduction number/rate, case fatality rate, transmission rate etc. As the focus of this paper is a meta-analysis of reproduction rate and case fatality rate, we extend our search of articles that solved for the estimation of reproduction rate and case fatality rate. We intend to look for articles in which reproduction number/rate or case fatality rate is estimated or calculated using some methods or methodology. Making our search more extensive, for reproduction number/rate, we searched on the basis of terms ‘coronavirus reproduction estimation’ in All Fields AND ‘COVID-19’ in the field Title/Abstract. This search in PubMed MEDLINE gives output for 29 articles to explore the parameter estimates of reproduction rates as well as the methodologies and epidemiological models. We reviewed title, keywords, abstracts and data and methods or methods/methodology, and references in these articles. We found three research articles which were based on a descriptive systematic review or general discussion focused on reproduction rates. Another two research articles were based on a systematic review, and only one research article was based on a meta-analysis of R_0_. Out of 29 articles, we found 24 research articles that provided 30 counts of R_0_ values with related statistics for a meta-analysis (Flowchart 1).

**Flowchart 1:**
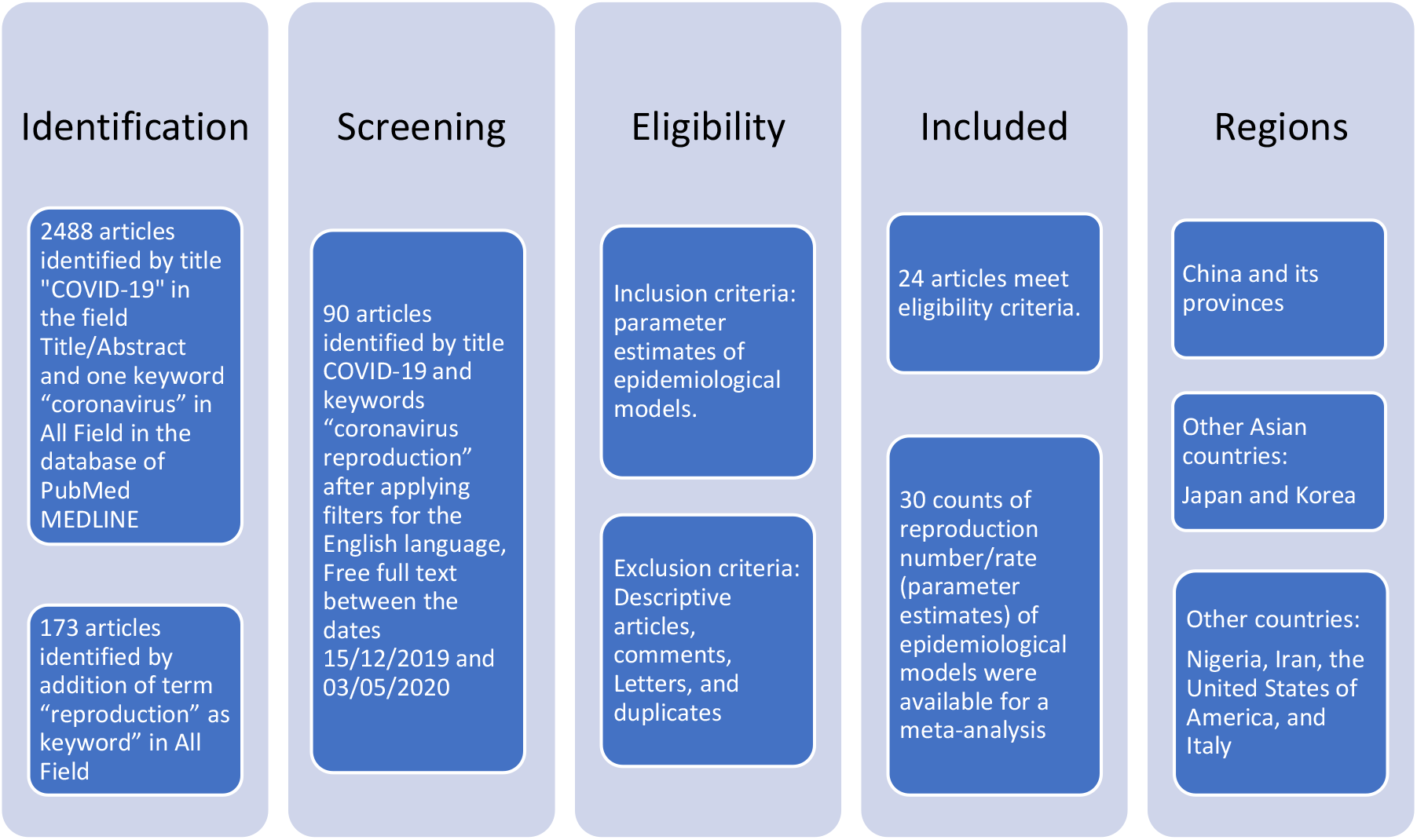
Flow diagram for the selected research articles for R_0_

### A systematic review of CFR

Similarly, for case fatality rate, considering 2488 numbers of research articles as the starting point of a systematic review, we searched with terms ‘coronavirus and mortality’ in All Fields AND ‘COVID-19’ in Title/Abstract that gave an output of 118 research articles, and with terms ‘fatality’ in All Fields AND ‘COVID-19’ in the Title/Abstract gave an output of 220 articles, and with terms, ‘fatality estimation risk’ OR ‘fatality estimation model’ OR ‘coronavirus mortality model parameter’ in All Fields AND ‘COVID-19’ in the Title/Abstract gave an output of 35 articles. Then, we applied the filters of the English language, Free full text, and between the dates 15/12/2019 and 03/05/2020, which gave an output of 24 articles. We reviewed these research articles searching for titles, abstract and keywords, methodology and references. Out of these 24 articles, only 17 qualified for quantifying parameter values of CFR (Flowchart 2). These 17 articles provided 29 counts of CFR for a meta-analysis.

**Flowchart 2:**
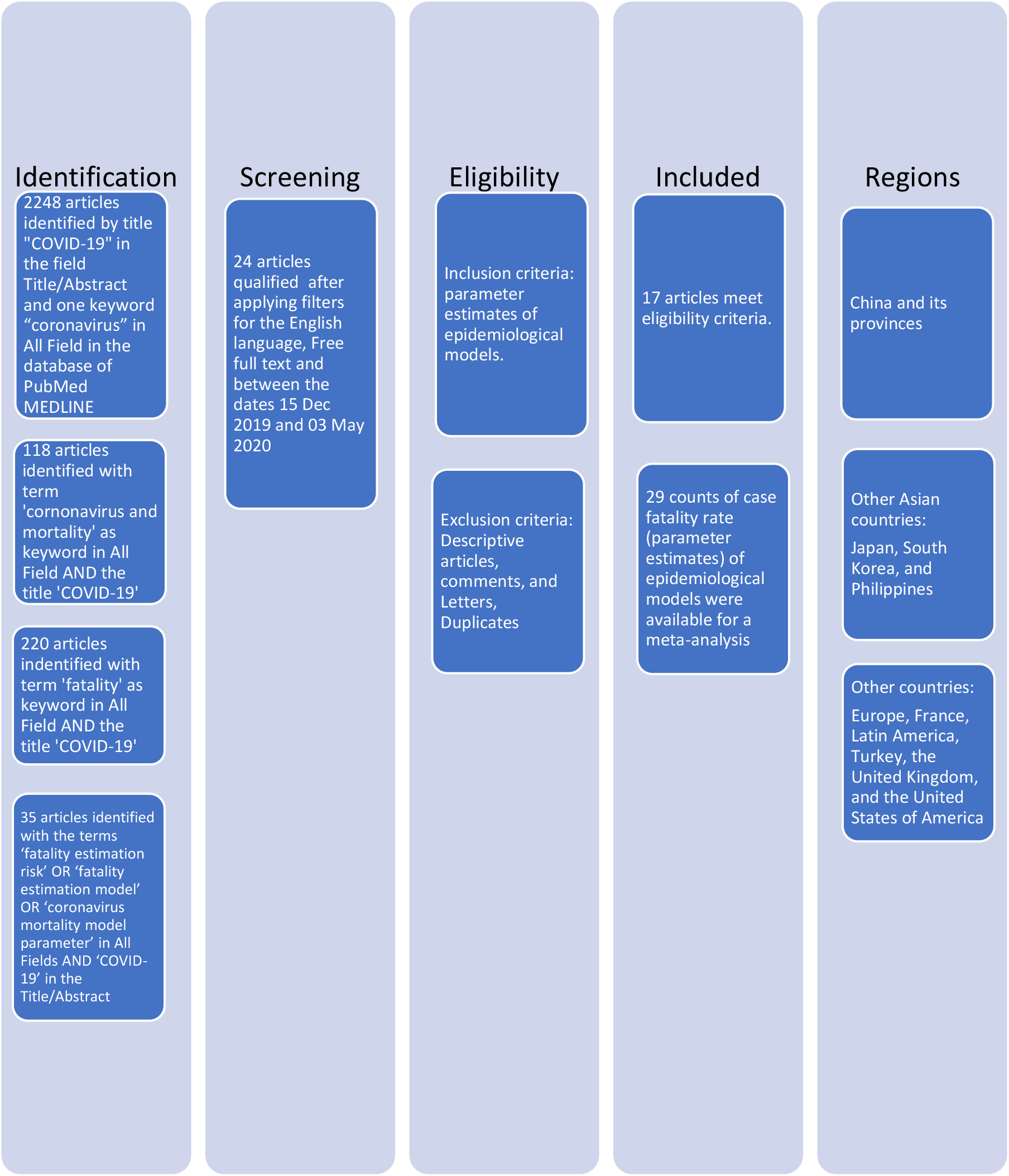
Flow diagram for the selected research articles for CFR

### Reproduction number/rate (R_0_) and Case fatality rate: parameter values of an epidemiological model

The review of methodologies of these 24 peer-reviewed articles for R_0_ and 17 peer-reviewed articles for CFR revealed that the studies lacked the control groups. The R_0_ and CFR values were retrieved from these published articles after careful reading and analysing the sections of ‘Data and Methods’ or ‘Methods’ or ‘Methodology’ and the ‘Results with tables and figures.’ This allowed to extract the date of publication, the period of study, application of methods, confirmed cases, susceptible cases, the R_0_ values, the CFR values, and relevant statistics such as confidence interval, standard error, sample size etc. For every statistic collected, we checked for any references mentioned against the source of statistic to avoid the duplication of estimates in two or more pieces of literature quoted from the same source of the work of literature. In this manner, the statistics of R_0_ and CFR computed in these studies qualified for a meta-analysis. The extracted R_0_ values were furthermore examined by the period of study. Many studies have given an effective reproduction rate (R_t_) along with R_0_ values; nevertheless, we consider the initial period of analysis and R_0_ values from the first phase of evolving epidemiology of the disease. A few of the studies have given several R_0_ values starting from the onset of the diseases until the end of the study. In such cases, we included the R_0_ values of the very onset period of the disease. The initial period or the first phase in these studies ranged between ∼5 days to ∼12 days. Analysing CFR, we included only adjusted case fatality rate standardised for age and sex distributions or for censored cases. Crude case fatality rates were excluded for a meta-analysis. Although several values of CFR were shown in a study, we included only those estimates based on deaths that comprise the sample of an epidemiological model. Two or more estimates of R_0_ and CFR were considered from the same study if it belonged to different regions or periods or were estimated from different method(s).

## Data extraction

We extracted the following variables: author, publication date, year, co-authors, sample size, mean statistics of R_0_, confidence interval, methods applied, standard deviation, and any other relevant statistics. All the studies, 24 for R_0_ and 17 for CFR were retained in the present study. These selected studies provided 30 counts of R_0_ and 29 counts of CFR for a meta-analysis. Table 1 and Table 2 show respectively for R_0_ and CFR, from the list of works of literature, the estimates of published R_0_ and CFR by authors applying different methodologies.

**Table 1:** Characteristics of included studies in the meta-analysis of R_0_.

| Author/<br>Study | Region/<br>Time period<br>(Date/Month/Y<br>ear) | Methodology | Basic<br>Reproduct<br>ion<br>Number/r<br>ate ($R_0$) | 95%<br>Confidence<br>Interval<br>(CI) | Confirmed<br>(C) and<br>Susceptible<br>(S) cases |
| --- | --- | --- | --- | --- | --- |
| Adegboye<br>et al. 2020 | Nigeria<br>27/02/2020-19/03/2020 | Bayesian method<br>(Short duration) | 4.98 | 2.65-8.41 | 318(C) |
| Adegboye<br>et al. 2020 | Nigeria<br>27/02/2020-11/04/2020 | Bayesian method<br>(Long duration) | 1.42 | 1.26-1.58 | 318(C) |
| Anastasso<br>poulou et<br>al. 2020 | Hubei Province<br>(China)<br>11/01/2020-20/01/2020 | Susceptible,<br>Infected,<br>Recovered and<br>Dead (SIRD)<br>models<br>(Long duration) | 7.09 | 5.84-8.35 | 59000000<br>(S) |
| Anastasso<br>poulou et<br>al. 2020 | Hubei Province<br>(China)<br>11/01/2020-16/01/2020 | Susceptible,<br>Infected,<br>Recovered and<br>Dead (SIRD)<br>models<br>(Short duration) | 4.8 | 3.36-6.67 | 59000000<br>(S) |
| Boldog et al. 2020 | Hubei Province (China)<br>23/01/2020-31/01/2020 | Time-dependent compartmental model, Galton–Watson branching process | 2.6 | 2.1-3.1 | - |
| Choi and Ki 2020 | Hubei Province (China)<br>20/01/2020-17/02/2020 | Susceptible (S), Exposed (E), Symptomatic Infectious (I) hospitalised (H) recovered or death (R): SEIHR model | 4.028 | 4.01-4.046 | 4992000 (S) |
| Choi and Ki 2020 | Korea<br>18/02/2020-24/02/2020 | Susceptible (S), Exposed (E), Symptomatic infectious (I) hospitalised (H) recovered or death (R): SEIHR model | 0.555 | 0.51-0.60 | 30 (C) |
| Jung et al. 2020 | China<br>08/12/2019-24/01/2020 | Delay distributions, Markov Chain Monte Carlo (MCMC) | 3.2 | 2.7-3.7 | 20 (C) |
| Kucharski et al. 2020 | Wuhan (China)<br>29/12/2019-23/01/2020 | Stochastic transmission dynamic model, Geometric random walk, Markov Chain Monte Carlo (MCMC) | 2.35 | 1.15-4.77 | - |
| Kuniya 2020 | Japan<br>15/01/2020-29/02/2020 | SEIR compartmental model | 2.6 | 2.4-2.8 | 239 (C) |
| Lai et al. 2020 | mainland China<br>31/12/2019-28/01/2020 | Pooling of estimates from different studies | 2.68 | 2.47-2.86 | 278 (C) |
| Muniz-Rodriguez et al. 2020 | Iran<br>19/02/2020-19/03/2020 | Generalised growth model | 4.4 | 3.9-4.9 | - |
| Muniz-Rodriguez et al. 2020 | Iran<br>19/02/2020-19/03/2020 | Epidemic doubling time | 3.5 | 1.3-8.1 | - |
| Peirlink et al. 2020 | USA<br>21/01/2020-04/04/2020 | SEIR compartmental model | 5.3 | 4.35-6.25 | 311357(C) |
| Riou and Althaus | Wuhan (China)<br>31/01/2020-29/01/2020 | Simulation | 2.2 | 1.4-3.8 | 5997 |
| Rocklov et al. 2020 | Japan<br>21/01/2020-19/02/2020 | Compartmental model using Susceptible, Infected, Infectious, and Recovered (SEIR) model | 14.8 | 5.3-19 | 619 (C) |
| Russo et al. 2020 | Lombardy (Italy)<br>05/01/2020-09/04/2020 | compartmental Susceptible/Exposed/ Infectious/ Recovered/ Dead (SEIRD) Model | 4.51 | 4.14-4.9 | 143626(C) |
| Sanche et al. 2020 | China CDC<br>15/01/2020-30/01/2020 | Hybrid deterministic–stochastic SEIR | 5.7 | 3.8-8.9 | 140 (C) |
| Shim et al. 2020 | South Korea<br>20/01/2020-06/03/2020 | empirical reporting delay distribution and simulating the generalised growth model | 1.5 | 1.4-1.6 | 6284(C) |
| Tang et al. 2020 | China<br>08/12/2019- | Departmental SEIHR model | 3.27 | 2.98-3.58 | 33 (C) |
|  | 04/01/2020 |  |  |  |  |
| Tsang and Bajpai 2020 | Wuhan<br>15/01/2020-03/03/2020 | Moments of Gamma distribution, Markov Chain Monte Carlo (MCMC) | 3.15 | 2.8-3.5 | - |
| Wang et al. 2020 | Hubei Province (China)<br>17/01/2020-08/02/2020 | Exponential growth (EG) method | 3.49 | 3.42-3.58 | - |
| Wang et al. 2020 | Hubei Province (China)<br>17/01/2020-08/02/2020 | Exponential growth (EG) method, Maximum likelihood estimation (ML), Sequential Bayesian method (SB) | 2.95 | 2.86-3.03 |  |
| Zhang et al. 2020 | Japan (Diamond Princess Cruise Ship- UK)<br>20/01/2020-17/02/2020 | Bootstrap sampling method | 2.28 | 2.06-2.52 | 355 (C)<br>3711 (S) |
| Zhao et al. 2020 | China<br>01/01/2020-15/01/2020 | exponential growth model (Long duration) | 2.56 | 2.49-2.63 | 2066(C) |
| Zhao et al. 2020 | China<br>10/01/2020-24/01/2020 | exponential growth model (Short duration) | 2.24 | 1.96-2.55 | - |
| Zhou et al. 2020 | China<br>10/01/2020-31/01/2020 | Dynamic compartmental model, Basic SEIR model | 5.316 |  | 118 (C)<br>11081000 (S) |
| Zhu et al. 2020 | China CDC<br>01/12/2019-23/01/2020 | MLE estimation, Poisson transmission<br>model | 2.54 | 2.49-2.6 | 3442 (C)<br>8348 (S) |
| Zhuang et<br>al. 2020 | Korea<br>20/01/2020-05/03/2020 | stochastic model | 2.6 | 2.3-2.9 | 6088(C) |
| Zhuang et<br>al. 2020 | Italy<br>06/02/2020-05/03/2020 | stochastic model | 3.2 | 2.9-3.5 | 3142(C) |
| **Overall Mean $R_0$ with 95% CI** |  |  | **3.72** | **3.43-4.04** | **30** |

Note: Unit of $R_0$ is expressed in persons.

**Table 2:** Characteristics of included studies in the meta-analysis for CFR.

| **Author/**<br>**Study** | **Region/**<br>**Period**<br>**(Date/Month/**<br>**Year)** | **Methodology** | **Case**<br>**fatality**<br>**rate**<br>**(CFR)** | **95%**<br>**Confidence**<br>**Interval**<br>**(CI)** | **Deaths/**<br>**Confirmed**<br>**cases** |
| --- | --- | --- | --- | --- | --- |
| Amariles et al. 2020 | Europe<br>06/03/2020-18/03/2020 | Stochastic differential evolution algorithm, SIRD model | 4.3 | 0.30-8.30 | 4664/88850 |
| Amariles et al. 2020 | Latin America<br>06/03/2020-18/03/2020 | Stochastic differential evolution algorithm, SIRD model | 1.95 | 0.80-3.10 | 14/1510 |
| Anastassopoulou et al. 2020 | Hubei Province (China)<br>11/01/2020-10/02/2020 | Susceptible, Infected, Recovered and Dead (SIRD) models | 2.94 | 2.9-3.0 | -/- |
| Bayham et al. 2020 | USA<br>15/02/2020-22/03/2020 | Infection mortality rate, MCKC simulations | 2.3 | 1.80-2.80 | - |
| Chatterjee et al. 2020 | Globally<br>01/12/2019-28/02/2020 | Descriptive: Adjusted fatality rate | 3.41 | 3.29-3.54 | 2859/83704 |
| Fu et al. 2020 | Overall<br>-- | Systematic review/Meta-analysis | 3.6 | 1.1- 7.2 | 43/- |
| Geldsetzer 2020 | USA | Rapid online Surveys/computer | 5 | 2.0-15.0 | -/1924 |
| Geldsetzer 2020 | UK | Rapid online Surveys/computer | 3 | 2.00-10.0 | -/1540 |
| Jung et al. 2020 | China (Fixed starting point)<br>08/12/2019-24/01/2020 | Delay distributions with fixed starting point, MCMC | 5.3 | 3.5-7.5 | 41/- |
| Jung et al. 2020 | China (Varying starting point)<br>13/01/2020-24/01/2020 | Delay distributions variable starting point, MCMC | 8.4 | 5.3-12.3 | 41/- |
| Kobayashi et al. 2020 | Hubei<br>31/12/2020-14/02/2020 | Descriptive: Adjusted fatality rate | 18 | 11-81 | - |
| Kobayashi et al. 2020 | Outside mainland China<br>31/12/2020-14/02/2020 | Descriptive: Adjusted fatality rate | 2.5 | 1-85 | - |
| Kobayashi et al. 2020 | Hubei<br>5 days | Descriptive: Infection fatality risk | 0.27 | 0.19- 0.38 |  |
| Michael Arieh P. Medina 2020 | Philippines<br>08/03/2020-06/04/2020 | simple linear regression model | 4.35 | 4.12-4.55 | 163/3000 |
| Mizumoto and Chowell 2020 | China<br>01/01/2020-11/02/2020 | Delay models, MCKC, Bayesian framework | 12.2 | 11.3-13.1 | Rolling/1117 |
| Öztoprak and Javed | Turkey<br>16/03/2020-31/03/2020 | linear regression analysis | 1.85 | 1.51-2.18 | 314/13531 |
| Öztoprak and Javed | France<br>16/03/2020-31/03/2020 | linear regression analysis | 1.979 | 1.79-2.15 | 48/2281 |
| Russell et al. 2020 | Japan<br>05/02/2020- | Age standardisation | 2.6 | 0.89-6.70 | 7/705 |
|  | 04/03/2020 |  |  |  |  |
| Shim et al. 2020 | South Korea<br>20/01/2020-26/02/2020 | Generalised growth model | 0.7 | 0.4-1.1 | 42/6284 |
| Verity et al. 2020 | China<br>01/01/2020-08/02/2020 | Bayesian Marko-Chain Monte Carlo (Adjusted for censoring) | 3.67 | 3.56- 3.8 | 24/70117 |
| Verity et al. 2020 | China<br>01/01/2020-08/02/2020 | Bayesian Marko-Chain Monte Carlo (Demographic adjustment) | 1.38 | 1.23- 1.53 | 24/70117 |
| Wang et al. 2020 | China<br>15/01/2020-11/03/2020 | binomial probability method | 3.9 | 3.80-4.10 | 3169/80793 |
| Wang et al. 2020 | China outside of Hubei<br>15/01/2020-11/03/2020 | binomial probability method | 0.87 | 0.72-1.00 | 3169/80793 |
| Wang et al. 2020 | China<br>15/01/2020-11/03/2020 | survival analysis method | 4.6 | 4.40-4.70 | 3169/80793 |
| Wang et al. 2020 | China outside of Hubei<br>15/01/2020-11/03/2020 | survival analysis method | .92 | 0.76-1.10 | 3169/80793 |
| Wilson et al. 2020 | China<br>21/02/2020-05/03/2020 | Time-delay adjusted case-fatality risk | 3.5 | 3.35- 3.61 | 2624/75569 |
| Wilson et al. 2020 | 82 countries<br>21/02/2020-05/03/2020 | Time-delay adjusted case-fatality risk | 4.2 | 2.58-6.87 | 15/354 |
| Yang et al.<br>2020 | China<br>10/01/2020-03/02/2020 | Linear<br>regression | 2.1 | 2.05-2.14 | -/- |
| Yang et al.<br>2020 | Hubei<br>10/01/2020-03/02/2020 | Linear<br>regression | 1.41 | 1.38-1.45 | -/- |
| **Overall Mean CFR (SD) with 95% CI** |  |  | **3.96** | **3.67-4.27** | **28** |

Note: Unit of CFR is expressed in per cent.

## Analyses

We performed the meta-analysis to estimate the mean effect size and its precision. The meta-analysis can be performed using the inverse variance method, fixed-effects model, and random-effects model. The Higgin’s & Thompson’s *I^2^* statistic, Tau-squared (*τ^2^*), and Cochran’s Q test were applied to test statistical heterogeneity among the selected studies. The test of heterogeneity was applied for understanding the application of the random-effects model versus the fixed-effects model in a meta-analysis. We plotted Forest plot using the random-effects model with prediction values of 95% confidence intervals. We plotted Funnel plot at 95%, 97.5%, and 99% confidence intervals to testify the publication bias in this meta-analysis of COVID-19.

## Results

### A meta-analysis of published R_0_ and CFR values

The average R_0_ value and CFR value was computed at a value of 3.72 (3.43–4.04) persons (Table 1) and 3.96 (3.67–4.27) per cent (Table 2), respectively, based on inverse variance method. We performed a test of heterogeneity to check whether these works of literature stem from the same population or from a universe of a population. The applied test of heterogeneity is shown in Table 3. The Higgin’s & Thompson’s *I^2^* statistic, which is the percentage of variability in the effect sizes not caused by sampling errors, greater than 75 per cent indicates a presence of high heterogeneity among these works of literature. The high value of 99.9% *I^2^* statistic confirms that these studies did not stem from the same population. The other two statistics which are tau-squared (*τ^2^*) statistic, the between-study variance in a meta-analysis, and Cochran’s Q-statistic, the difference between the observed effect sizes and fixed-effects model, are significant for R_0_ as well as for CFR (Table 3). These tests of heterogeneity were quite important for deciding the application of fixed-effects or random-effects models for a meta-analysis. The fixed-effects model assumes that the studies stem from the same population whereas the random-effects model is based on the fact the studies stem from the universe of population. Results from the test of heterogeneity suggest for applying the random-effects model rather than for applying the fixed-effects model for a meta-analysis. We computed the mean R_0_ value and mean CFR value (mean effect sizes) were computed, using the random-effects model in a meta-analysis.

**Table 3:** Test of heterogeneity for sample in a meta-analysis.

| Test of Heterogeneity | $R_0$ | CFR |
| --- | --- | --- |
| $I^2$ statistic | 99.9%<br>(99.9% - 99.9%) | 99.7%<br>(99.6% - 99.7%) |
| Tau-squared ($\tau^2$) | 2.42<br>(0.84-4.75) | 1.22<br>(1.06- 5.81) |
| Cochran's Q statistics | 22701.5* | 7657.4* |
| **n** | 30 | 28 |

Source: Own calculations
\*: significant at 95% of confidence interval

The estimates of R_0_ and CFR were computed using the random-effects model based on data shown in Table 1 and Table 2. We have summarised the estimates of R_0_ in Table 4 and CFR in Table 5 from meta-analysis, using on all data and excluding outliers. We estimated mean value of R_0_ and CFR, after excluding outliers, at 3.11 (2.49–3.71) persons (Table 4: column (f)) and 2.63 (2.18–3.08) per cent (Table 5: column (f)), respectively, based on the pieces of evidence available across the countries. These estimates are accounted for heterogeneity among the studies and lies within a narrow confidence interval. Hence, these estimates qualify as a high precision estimate. The estimated R_0_ value based on the random-effects model is slightly lower than that of based on the inverse variance method.

**Table 4:**
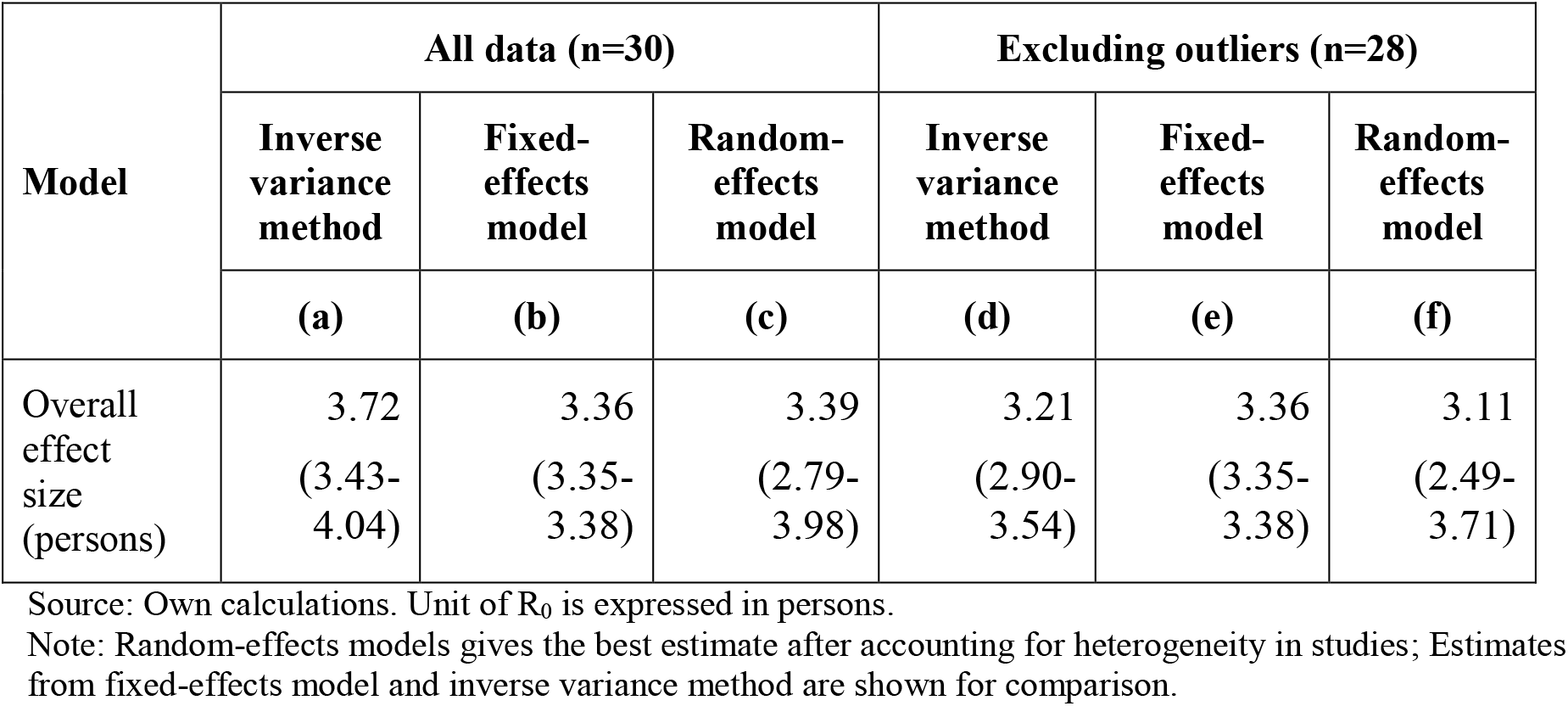
Overall effect size based on various methods for R_0_.

| Model | All data (n=30) |  |  | Excluding outliers (n=28) |  |  |
| --- | --- | --- | --- | --- | --- | --- |
|  | Inverse variance method | Fixed-effects model | Random-effects model | Inverse variance method | Fixed-effects model | Random-effects model |
|  | (a) | (b) | (c) | (d) | (e) | (f) |
| Overall effect size (persons) | 3.72<br>(3.43-4.04) | 3.36<br>(3.35-3.38) | 3.39<br>(2.79-3.98) | 3.21<br>(2.90-3.54) | 3.36<br>(3.35-3.38) | 3.11<br>(2.49-3.71) |

Source: Own calculations. Unit of $R_0$ is expressed in persons.
Note: Random-effects models gives the best estimate after accounting for heterogeneity in studies; Estimates from fixed-effects model and inverse variance method are shown for comparison.

**Table 5:** Overall effect size based on various methods for CFR.

| Model | All data (n=28) |  |  | Excluding outliers (n=24) |  |  |
| --- | --- | --- | --- | --- | --- | --- |
|  | Inverse variance method | Fixed-effects model | Random-effects model | Inverse variance method | Fixed-effects model | Random-effects model |
|  | (a) | (b) | (c) | (d) | (e) | (f) |
| Overall effect size (per cent) | 3.96<br>(3.67-4.27) | 2.17<br>(2.15-2.19) | 3.20<br>(2.76-3.64) | 2.79<br>(2.58-3.02) | 2.16<br>(2.14-2.18) | 2.63<br>(2.18-3.08) |

Source: Own calculations; Unit of CFR is expressed in per cent.
Note: Random-effects models gives the best estimate after accounting for heterogeneity in studies; Estimates from fixed-effects model and inverse variance method are shown for comparison.

Furthermore, for examining a regional variation, the meta-analysis was performed by subgroups of countries. A minimum sample of three studies is required for performing a subgroup analysis in a meta-analysis. The regional subgroups identified for meta-analysis of R_0_ are ‘China and its provinces’, ‘other Asian countries’ that includes studies based on data from Japan and Korea, and ‘other countries’ that includes studies based on data from Nigeria, Iran, Italy, and the United States of America (USA). Figure 1 and Figure 2 show Forest plot showing mean effect sizes by regional subgroups for R_0_ and CFR, respectively, along with the overall effect size, based on the random-effects model. The mean R_0_ values for the regional subgroup ‘China and its provinces’ was 3.21 (2.73–3.68) persons, and for ‘other Asian countries’ was 1.90 (1.06–2.74) persons and for ‘other countries’ was 3.83 (2.44–5.22) persons (Table 6). The test of regional subgroup differences using random-effects model is significant with a p-value of 0.013. This confirms that the estimated mean R_0_ values are significantly different across these regions. The results revealed that, among these regions, it is the highest for the subgroup ‘other countries’, wherein it is the highest for the USA. The regional subgroups identified for CFR meta-analysis are ‘China and its provinces’, ‘other Asian countries’ that includes study based on data from Japan, South Korea, and the Philippines, and ‘other countries’ that includes studies based on data from Europe, France, Latin America, Turkey, the United Kingdom (UK), and the USA. The mean CFR values for ‘China and its provinces’ was 2.53 (1.91–3.14) per cent, for ‘other Asian countries’ was 2.56 (−0.26–5.38) per cent, and for ‘other countries’ was 2.78 (2.08–3.47) per cent (Table 7). The test for regional subgroup differences using the random-effects model was not significant (p = 0.865). Hence, it implies that the CFR did not vary significantly across these regions despite the fact that the CFR was the highest in the USA and the lowest in South Korea.

**Table 6:**
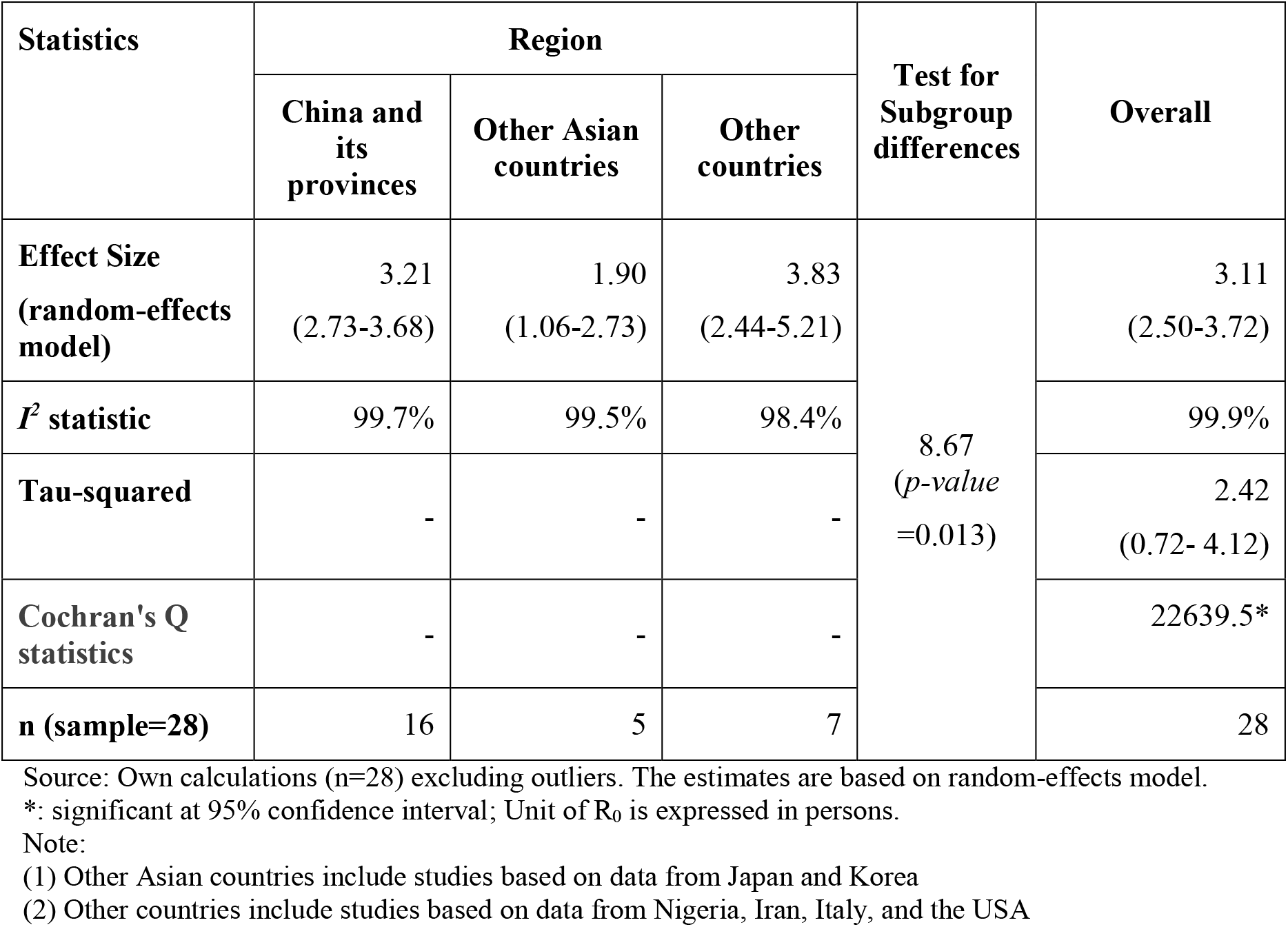
Effect size and test of heterogeneity for R_0_ by region and overall.

| Statistics | Region |  |  | Test for Subgroup differences | Overall |
| --- | --- | --- | --- | --- | --- |
|  | China and its provinces | Other Asian countries | Other countries |  |  |
| **Effect Size (random-effects model)** | 3.21<br>(2.73-3.68) | 1.90<br>(1.06-2.73) | 3.83<br>(2.44-5.21) | 8.67<br>(*p-value* =0.013) | 3.11<br>(2.50-3.72) |
| **$I^2$ statistic** | 99.7% | 99.5% | 98.4% |  | 99.9% |
| **Tau-squared** | - | - | - |  | 2.42<br>(0.72- 4.12) |
| **Cochran's Q statistics** | - | - | - |  | 22639.5* |
| **n (sample=28)** | 16 | 5 | 7 |  | 28 |

Source: Own calculations (n=28) excluding outliers. The estimates are based on random-effects model.
\*: significant at 95% confidence interval; Unit of $R_0$ is expressed in persons.
Note:
(1) Other Asian countries include studies based on data from Japan and Korea
(2) Other countries include studies based on data from Nigeria, Iran, Italy, and the USA

**Table 7:** Effect size and test of heterogeneity for CFR by region and overall.

| Statistics | Region |  |  | Test for Subgroup differences | Overall |
| --- | --- | --- | --- | --- | --- |
|  | China and its provinces | Other Asian countries | Other countries |  |  |
| **Effect Size (random-effects model)** | 2.53<br>(1.91-3.14) | 2.55<br>(-0.37-5.46) | 2.77<br>(2.07-3.47) | 0.22<br>(*p-value* =0.868) | 2.63<br>(2.18-3.08) |
| **$I^2$ statistic** | 99.9% | 99.4% | 95.9% |  | 99.7% |
| **Tau-squared** | - | - | - |  | 1.06<br>(0.61- 3.48) |
| **Cochran's Q statistics** | - | - | - |  | 8060.72* |
| **n (sample=24)** | 11 | 3 | 10 |  | 24 |

Source: Own calculations (n=24) excluding outliers. The estimates are based on random-effects model.
\*: significant at 95% confidence interval; Unit of CFR is expressed in per cent.
Note:
(1) Other Asian countries include studies based on data from Japan, South Korea, and Philippines
(2) Other countries include studies based on data from Europe, France, Latin America, Turkey, the UK, and the USA

**Figure 1.**
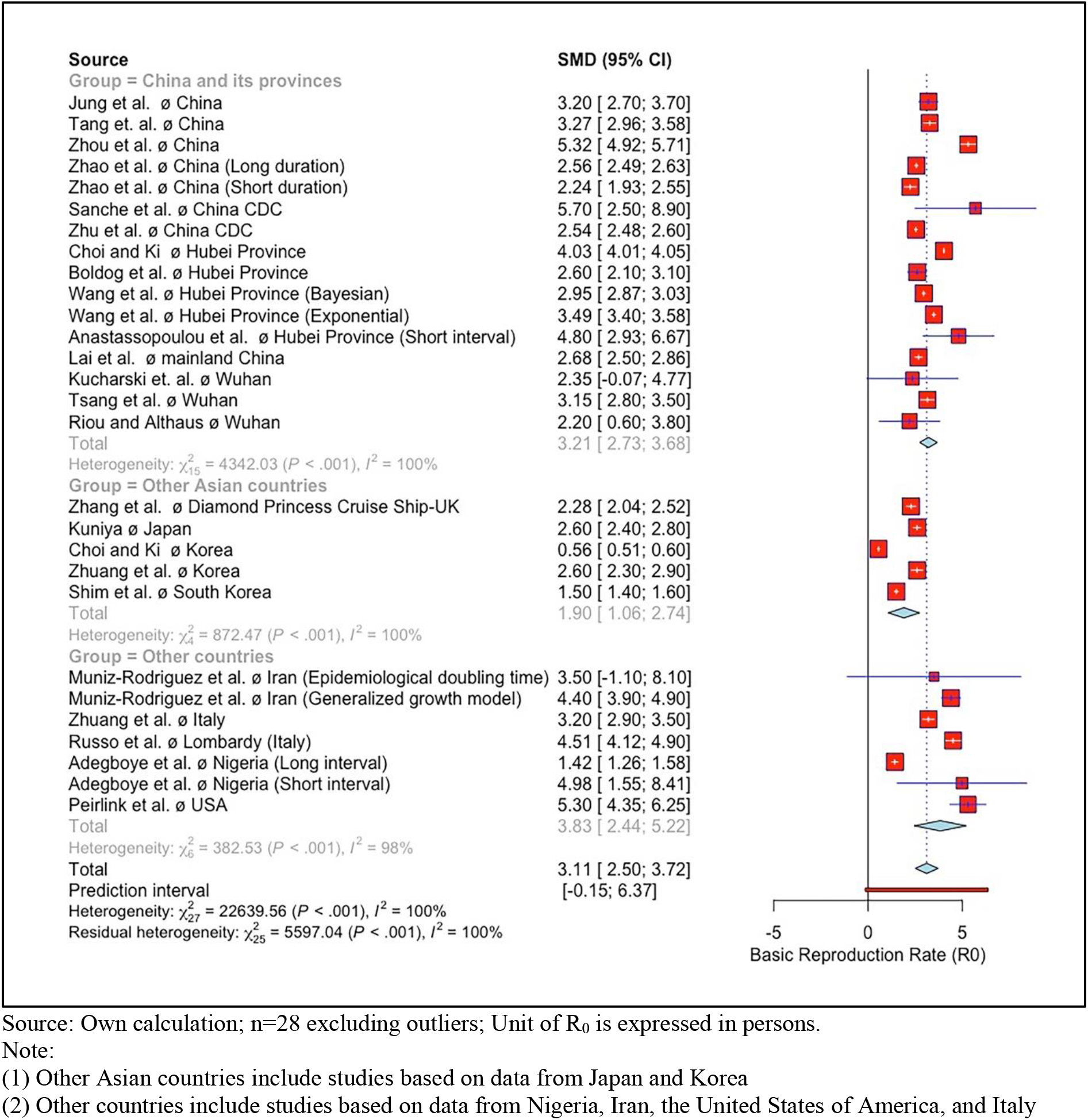
Forest plot of R_0_ values based on random-effects model, by regional subgroups

**Figure 2.**
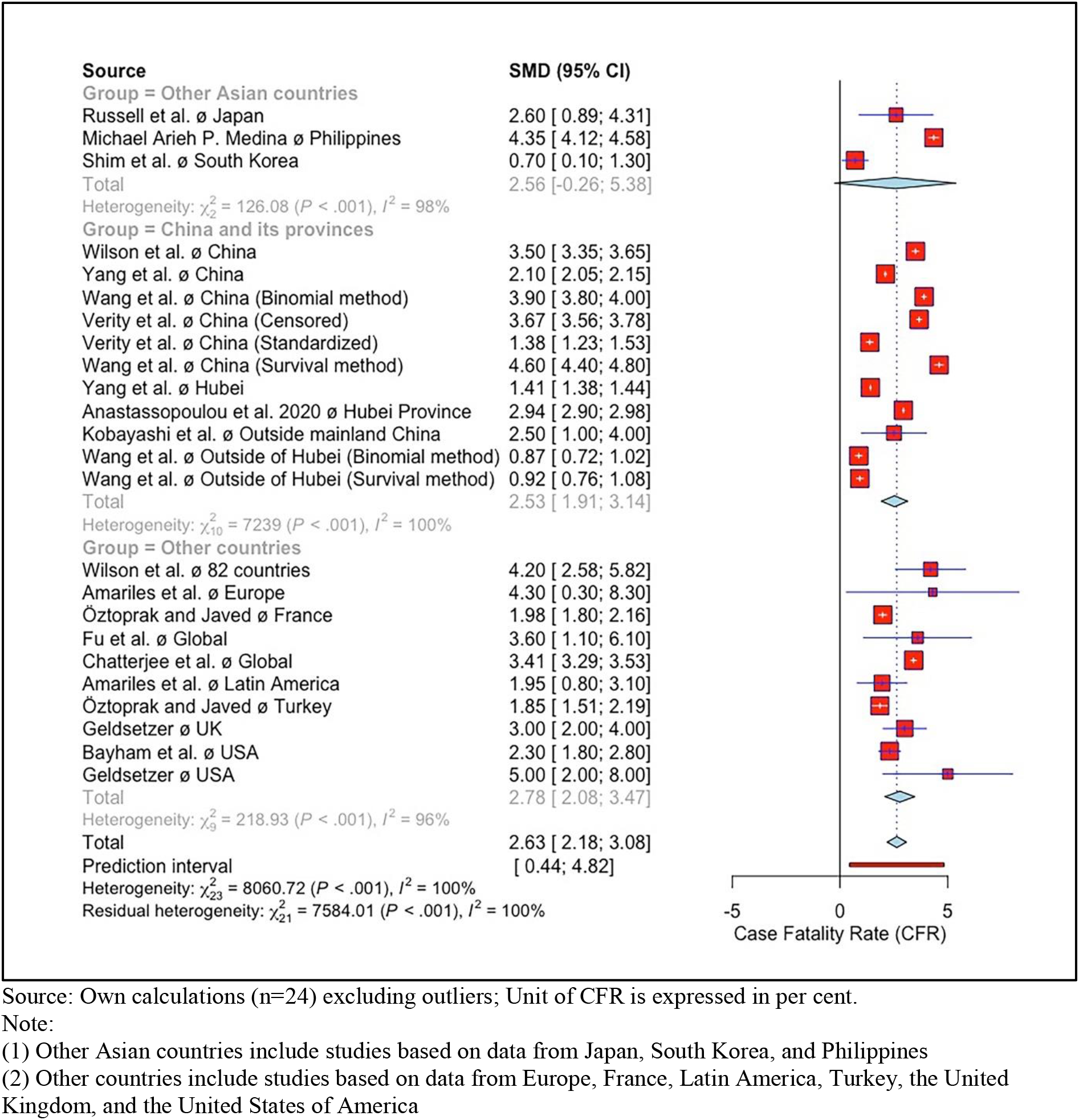
Forest plot of CFR values based on random-effects model, by regional subgroups

We have also looked at publication bias to test the significance of these studies for a meta-analysis. The Funnel plot for R_0_ values computed from the random-effects model is shown in Figure 3. This Funnel plot clearly shows that the selected studies of R_0_ for this meta-analysis are significant at one per cent level of significance, with the exception of two studies which are Muniz-Rodriguez et al. 2020 with the method of doubling time and Kucharski et al. 2020. Similarly, the Funnel plot of CFR is shown in Figure 4, based on the random-effects model. It clearly shows that all studies are significant at one per cent level of significance. The results from the Funnel confirm that even the small sample size studies have also been published in addition to moderate sample size studies and large sample sized studies which are generally project-based.

**Figure 3.**
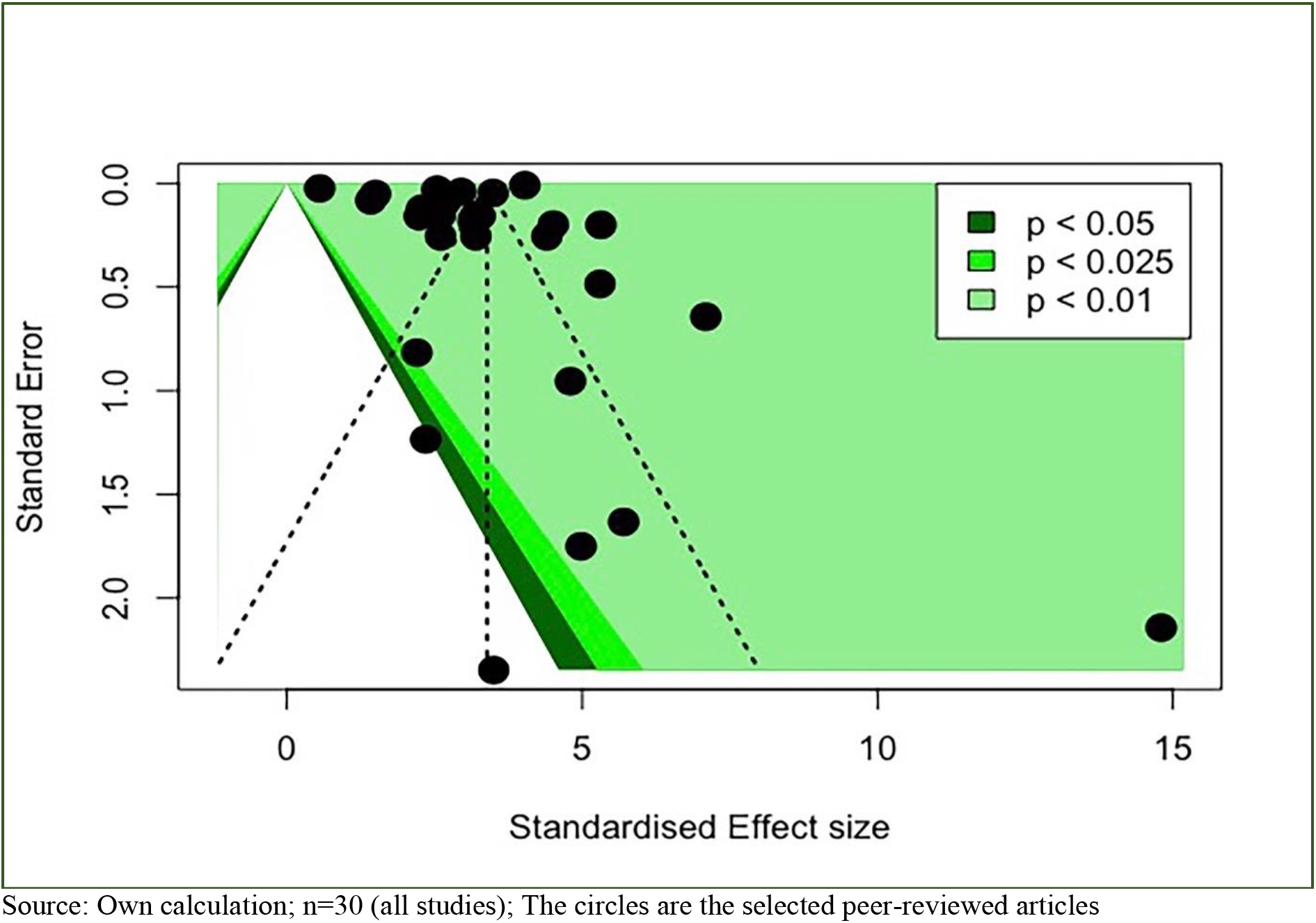
Funnel plot for R_0_ values based on random-effects model

**Figure 4.**
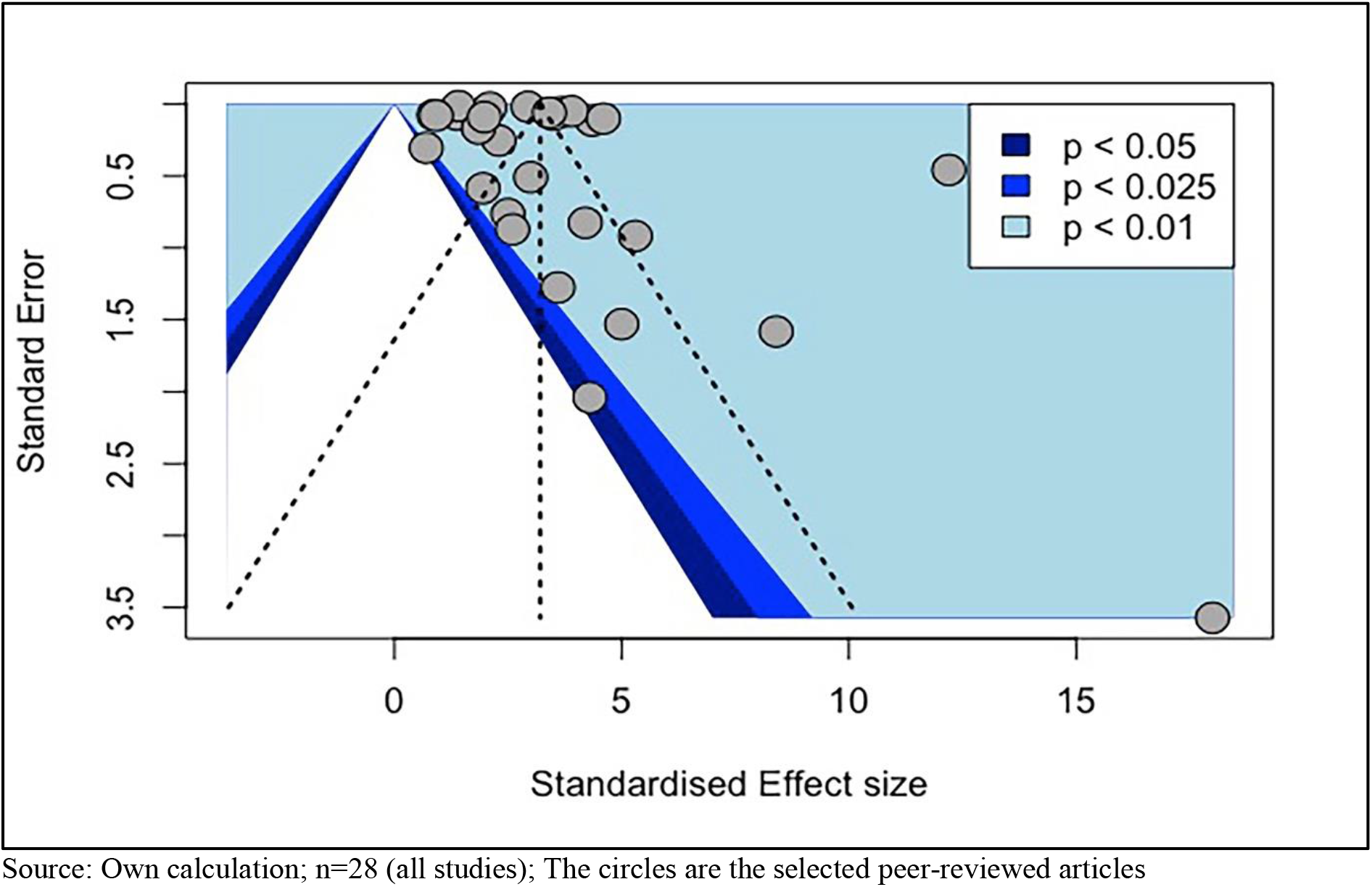
Funnel plot for CFR values based on random-effects model

## Discussion

This study aims to provide a summary statistic of the basic reproduction rate (R_0_) and the case fatality rate (CFR) for a generalised population based on peer-reviewed published estimates of R_0_ and CFR from epidemiological models applied on a susceptible population. After an electronic search for such conditions between the dates 15 December 2019 and 3 May 2020 and applying various inclusion and exclusion criteria, this study came across 24 and 17 works of literature for R_0_ and CFR, respectively that qualified for a meta-analysis. These studies provided 30 counts of R_0_ and 29 counts of CFR for a meta-analysis. The study examined the characteristics of studies and computed the overall effect size or mean R_0_ and CFR value. We applied the test of heterogeneity which are Higgin’s & Thompson’s *I^2^* statistic, tau-squared (*I^2^*) statistic, and Cochran’s Q-statistic. These test of heterogeneity reveals a high heterogeneity across the studies. The studies included in the meta-analysis had the sources of errors not only from sampling errors but also from the distributions of individual mean effect size. The R_0_ and CFR values extracted from different studies had their own distributions that were different from an overarching distribution. Therefore, the random-effects model was most appropriate for computing mean R_0_ value and mean CFR value based on a meta-analysis. After excluding outliers, the estimate of R_0_ and CFR from these studies was calculated at 3.11 (2.49–3.71) persons (Table 4: column (f)) and 2.63 (2.18–3.08) per cent (Table 5: column (f)), respectively, based on the random-effects model. For R_0_, the variation by subgroups of regions was significant. It reveals that the R_0_ values were significantly different across these regional subgroups. However, the regional subgroups differences for CFR was not a significant one. Hence, we conclude that the CFR did not vary across the regions.

Acknowledging the severity of this disease, the estimated R_0_ value of 3.11 persons and in a narrow confidence interval is a higher and riskier statistic applicable for any generalised population. This R_0_ statistic implies that one infectious person is transmitting to three other susceptible persons in the absence of any control measures. The R_0_ value is overall effect size based on various heterogeneous studies. Therefore, this R_0_ value is most reasonable and sensible for a country or a region encountering the emergence of COVID-19 during the first phase or in the very initial stage of this infectious disease. The meta-analysis by regional subgroups reveals the variation in R_0_ across the regions (Table 6). Therefore, this study suggests that R_0_ values based on a meta-analysis from the pieces of evidence across the regions would range between 1.90 (1.06–2.73) person and 3.83 (2.33–5.21) persons. The CFR statistic is based on the period of approximately one-and-a-half months, but it translates to toll deaths in a short span of time. The results of regional subgroup analysis in meta-analysis confirm that the CFR did not vary across the regions. The estimated CFR value of 2.63 per cent (Table 7) without much variation is applicable to any generalised population. For a developing country like India and the second most populated country in the world, the CFR value of 2.63 is most dreadful. Necessary precautions and strategies are utmost important as early as possible to prevent the outbreak of this disease.

## Conclusion

This paper suggests a robust estimate of R_0_ which is 3.11 (2.49–3.71) persons in the absence of any control measures and a robust estimate of CFR equals to 2.63 (2.18–3.08) per cent for a generalised population in a period of one-and-a-half months from the onset of disease COVID-19. The analysis by subgroups of regions using the Forest plot confirms a significant variation for R_0_, but the same is not found significant for CFR. The R_0_ values would most probably be in the range of 1.90 (1.06–2.73) person and 3.83 (2.33–5.21) persons for a region. The Funnel plot confirms that the included studies were significant, and therefore, it establishes the robustness of R_0_ and CFR based on the data of these studies in a meta-analysis. We proclaim that one person is likely to infect three persons in the absence of any control measures and the around three per cent of the population are at the risk of death in a period of one-and-a-half months from the onset of disease COVID-19 in a generalised population.

The estimates of R_0_ and CFR are unequivocally applicable to any generalised population at the point of emergence of the disease COVID-19. Hence these estimates are worthwhile for a region/country and its lower geography. These robust estimates are applicable for developing country India and its states or districts.

### Limitation of the study

This study is based on a meta-analysis of recently published articles that estimated for parameters of epidemiological models for COVID-19. The period for analysis for COVID-19 is more than three months from 15 December 2019 to 3 May 2020. We retrieved studies which are peer-reviewed research papers and are mostly from the regions which have encountered the epidemic in an area or pandemic in a country or nation at the very early emergence of SARSCoV-2. Many of these peer-reviewed studies have used data mainly from China and its provinces. Some of these studies had analysed data from other Asian countries and a few from other parts of the world. Therefore, a wide and rich regional view of data was not available in the period of study. Most of these studies in itself have the disadvantage of small sample size and missing information on the time of onset of SARS-CoV-2. Accordingly, the authors of these selected published papers have used generalised epidemiological models to get robust estimates. Most of these have used time-varying models using simulation methods and the moments of statistical distributions for estimating parameters of the epidemiological models. We also, in order to overcome such limitations of small sample size, have estimated the mean R_0_ and CFR values using the random-effects model which make the estimation of parameter based on the assumption that these studies stem from a universe of population. Therefore, the estimates of R_0_ and CFR in this study is robust and applicable to a generalised population. In addition to that, nonetheless, the Funnel plot for both R_0_ and CFR showed that these publications are important to consider for a meta-analysis, as these studies are found statistically significant for examining COVID-19.

## Data Availability

This Article is based on Published Literature search through PubMed MEDLINE, Endnote and Google Scholar.

## Acknowledgments

*Authors would like to thank Prof. K.S. James, Director and Senior Professor, International Institute for Population Sciences (IIPS), Mumbai, for his recommendations and suggestions to carry out this study and for providing constructive comments which helped greatly in improving the quality of the policy brief. IIPS team of researchers on Estimation and Projection of COVID 19 Cases would also like to thank him for initiating policy briefs on COVID-19*.

## IIPS team of researchers on Estimation and Projection of COVID 19 Cases

Dr. Sayeed Unisa, Professor and Head, Department of Mathematical Demography and Statistics International Institute for Population Sciences, Mumbai. (email id)

Dr. Chander Shekhar, Professor, Department of Fertility Studies, International Institute for Population Sciences, Mumbai. (email id)

Dr. Usha Ram, Professor, Department of Department of Public Health & Mortality Studies International Institute for Population Sciences, Mumbai. (email id)

Dr. Laxmi Kant Dwivedi, Assistant Professor, Department of Mathematical Demography and Statistics, International Institute for Population Sciences, Mumbai. (email id)

Dr. Suryakant Yadav, Assistant Professor, Department of Development Studies, International Institute for Population Sciences, Mumbai. (email id)

Dr. Preeti Dhillon, Assistant Professor, Department of Mathematical Demography and Statistics, International Institute for Population Sciences, Mumbai. (email id)

Mr. Pawan Kumar Yadav, Research Scholar, Mumbai, India

M. Kishore, M.Phil. Research Scholar, International Institute for Population Sciences, Mumbai. (email id)

## Notes

### Competing Interest Statement

The authors have declared no competing interest.

### Clinical Trial

This study is based on published Literatures so it is not registered for clinical trail.

